# The Impact of Post-Acute Care on Depression and Anxiety in Stroke Patients: A Prospective Study to Explore the Mediating Effect of Cognitive Function

**DOI:** 10.1101/2023.07.13.23292636

**Authors:** Shi-Jer Lou, Hsiu-Fen Lin, Yu-Tsz Shiu, Hon-Yi Shi

## Abstract

**BACKGROUND:** Cognitive function was significantly associated with post-stroke depression and anxiety in stroke patients. However, no studies have examined whether there is an interaction. this study purposed to investigate whether cognitive function mediates the effect of enrollment in post-acute care (PAC) programs on depression or anxiety in stroke patients and whether the indicators are moderated in the pathway.

**METHODS:** This is a prospective observational cohort study. A group of patients who had received PAC for stroke at one of two medical centers (PAC group, n = 2,087) was compared with a group who had received standard care for stroke at one of four hospitals (three regional hospitals and one district hospital; non-PAC group, n = 1,591) in Taiwan from March, 2015, to March, 2022. The effects of PAC on cognitive function and depression and anxiety at baseline, 12^th^ week, and 1^st^ year after rehabilitation were investigated using structural equation modeling (SEM). The effect of each variable on the moderation of different pathways in the model was analyzed using AMOS 23.0, and The SPSS PROCESS macro also was used to perform mediation analysis.

**RESULTS:** The PAC program had a mediating effect on cognition and depression at week 12 (a*b= -0.098, P<0.05) on cognition and anxiety at week 12 (a*b= -0.107, P<0.05), and the PAC program had a direct effect on depression and anxiety in the first year. It was found that acute lengths of stay had a significant moderation effect in the model (X*W→M=0.204, P=0.002), but the model lost its mediating effect when the moderation variable was added.

**CONCLUSIONS:** Patients with stroke should receive post-acute care as soon as possible to improve their cognitive function after rehabilitation, to maximize the effectiveness of treatment for mental disorders, and to reduce the burden of the disease.

**WHAT IS KNOWN:**

- Research suggests that cognitive function, including depression and anxiety, significantly improved for patients using post-acute care (PAC).
- Cognitive function was significantly associated with post-stroke depression and anxiety in patients with stroke.

**WHAT THE STUDY ADDS:**

- PAC had a mediating effect on cognition and depression at week 12 on cognition and anxiety at week 12, and it also had a direct effect on depression and anxiety in the first year.
- Acute lengths of stay had a significant moderation effect in the model, but the model lost its mediating effect when the moderation variable was added.
- Patients with stroke should receive post-acute care as soon as possible to improve their cognitive function after rehabilitation, to maximize the effectiveness of treatment for mental disorders, and to reduce the burden of the disease.

## INTRODUCTION

The post-acute care (PAC) program can significantly reduce disability and death in patients with stroke.^1, 2^ The National Health Insurance Agency announced the PAC program in 2014, designed to replace long-term hospitalization in medical centers.^3^ It allows patients to continue to receive appropriate care after treating acute illness. Initially, the PAC program was established for acute stroke patients who are medically stable with mild to moderate to severe functional impairment and who have the potential for active rehabilitation. After PAC was implemented, it was found that the patients receiving post-acute care showed improvement in all indicators, including activities of daily living, mobility, swallowing, and cognitive function.^4–6^

Research suggests that cognitive function, including depression and anxiety, significantly improved for patients using PAC.^7, 8^ Moreover, cognitive function was significantly associated with post-stroke depression and anxiety in stroke patients.^9, 10^ However, no studies have examined whether there is an interaction. Since depression and anxiety in patients with stroke vary over time, research must examine the mediating and moderating effects at different time points to gain a more detailed understanding of the onset of depression and anxiety symptoms in stroke patients after PAC rehabilitation. Therefore, the present study investigated whether cognitive functioning affected depression and anxiety in stroke patients in post-acute care and whether demographic, clinical, and quality-of-care characteristics had a moderating effect on the model.

## METHODS

### Study Design and Sample

This study prospectively recruited 3,678 stroke patients who received rehabilitation program at two medical centers, three regional hospitals, and one district hospital in southern Taiwan from March 1, 2014 to March 1, 2022. If patients were eligible for the PAC program and agreed to be referred to a district or regional hospital for PAC, they were selected for the PAC group. If the patients continued to receive acute care or were discharged from the medical center, they were selected for the non-PAC group. The patients’ physical functional status at the time of admission, at week 12, and at year one was tracked using questionnaires administered by trained research assistants.

These inclusion criteria were as follows: (1) Patients with acute cerebrovascular disease who were medically stable (controllable vital characteristics within 72 hours); (2) Patients with functional status assessed as mild to moderate or severe functional impairment (Modified Rankin Scale, mRS 2-4);^11^ (3) Patients with active participation in rehabilitation and the potential for positive rehabilitation. The exclusion criteria included patients who were under 18 with cognitive impairment (Mini-Mental State Examination, MMSE=0)^12^ and the inability to communicate. The study protocol was approved by the Institutional Review Board of the Kaohsiung Medical University Hospital (KMUH-IRB-20140308).

### Research Instruments

The medical record review included three main attributes: demographic attributes, clinical attributes, and healthcare quality attributes. The mRS assesses the degree of dependence on daily living activities in stroke patients and is divided into seven levels, with 0 representing no symptoms, 5 representing severe disability, and 6 representing death. The MMSE is an 11-question scale measuring time and place orientation, attention and computation, immediate and short-term memory, language, and visuospatial abilities. The maximum score is 30, with higher scores indicating better ability and scores below 25 indicating severe (≤9), moderate (10-20), or mild (21-24) impairment, respectively. The Beck Depression Inventory (BDI) comprises 21 questions, each with four options, ranging from 0 (not at all) to 3 (severe).^13^ The total score is 63, with higher scores indicating more severe depression. The Beck Anxiety Inventory (BAI) comprises 21 questions, each with four options, ranging from 0 (not at all) to 3 (severe).^14^ Its total score is 63, with higher scores indicating more severe anxiety.

### Research Variables

The independent variable was participating in PAC program or not. The dependent variables were the BDI and BAI scores. The mediating variable was the MMSE score. The moderating variables were age, gender, education, body mass index (BMI), smoking, alcohol drinking, acute lengths of stay, readmission in 30 days, stroke recurrence, use of a urinary catheter or nasogastric tube, hypertension, diabetes mellitus, hyperlipidemia, atrial fibrillation, and previous stroke.

### Statistical Analysis

The unit of analysis in this study was the individual patient with stroke. This study weighted the independent and moderating variables with the inverse probability of treatment weighting (IPTW) to balance the differences in the relevant variables between the two groups.

We used the structural equation model (SEM) to determine the mediating effects, explain the path coefficients, and investigate the mediating effects at different time points. We also used the cross-delay model with PAC as the independent variable, cognitive function at different time points as the mediating variable, and depression symptom and anxiety symptom at different time points as the dependent variables.

The mediating and moderating effect analysis focused on the critical moderators, and the model with the most mediating effect in SEM was used to explore the moderating variables.

Demographic, clinical, and quality of care characteristics were used to find the most moderating variables, which moderated the path between the independent and mediating variables and between the mediating and dependent variables. Therefore, the mediating and moderating effect model was built accordingly.

We performed statistical analysis using AMOS 23.0 and SPSS 21.0 software. The PROCESS module provided by Hayes (2013) for SPSS syntax was installed in SPSS. The models (Model 7) (Model 14) and (Model 58) were also used to elaborate the notion of mediation and interference and to analyze the mixed models.

## RESULTS

### Before and After IPTW

2,087 patients were in the PAC group with a mean age of 68.41 years (standard deviation, SD 13.02 years), and the majority were male patients (1,345, 64.4%); 1,591 patients were in the non-PAC group with a mean age of 70.12 years (SD 13.89 years), and the majority were male patients (1,011, 63.5%). They were weighted by IPTW and the distribution of the variables in the two groups was not significantly different (Table 1).

**Table 1.** Baseline Characteristics of the Study Population Before and After Inverse Probability of Treatment Weighting (IPTW) (N=3,678)

| Characteristics |  | Before IPTW |  |  | After IPTW |  |  |
| --- | --- | --- | --- | --- | --- | --- | --- |
|  |  | Non-PAC(n=1,591) | PAC (n=2,087) | P value | Non-PAC | PAC | P value |
| Age, years |  | 70.12±13.89 | 68.41±13.02 | 0.168 | 69.93±14.28 | 69.36±12.59 | 0.741 |
| Gender | Male | 1,011(63.5%) | 1,345(64.4%) | 0.848 | 291 | 300 | 0.693 |
|  | Female | 580(36.5%) | 742(35.6%) |  | 178 | 174 |  |
| Education, years |  | 8.54±5 | 8.02±1.77 | 0.114 | 8.11±5.05 | 7.98±1.79 | 0.593 |
| Body mass index, kg/m<sup>2</sup> |  | 24.29±3.42 | 24.32±4.01 | 0.918 | 24.48±3.61 | 24.37±3.95 | 0.673 |
| Acute lengths of stay, days |  | 15.56±9.88 | 18.71±9.55 | <0.001 | 17.07±11.01 | 17.22±9.24 | 0.815 |
| Readmission in 30 days | Yes | 176(11.1%) | 107(5.1%) | 0.016 | 38 | 34 | 0.591 |
|  | No | 1,415(88.9%) | 1,980(94.9%) |  | 430 | 439 |  |
| Stroke recurrence | Yes | 217(13.6%) | 55(2.6%) | <0.001 | 35 | 31 | 0.573 |
|  | No | 1,374(86.4%) | 2,032(97.4%) |  | 433 | 443 |  |
| Nasogastric tube | Yes | 449(28.2%) | 456(21.8%) | 0.114 | 125 | 119 | 0.587 |
|  | No | 1,142(71.8%) | 1,631(78.2%) |  | 354 | 343 |  |
| Foley catheter | Yes | 279(17.5%) | 154(7.4%) | 0.001 | 57 | 53 | 0.650 |
|  | No | 1,312(82.5%) | 1,933(92.6%) |  | 412 | 420 |  |
| Hypertension | Yes | 1,129(71.0%) | 1,559(74.7%) | 0.337 | 347 | 352 | 0.924 |
|  | No | 462(29.0%) | 528(25.3%) |  | 121 | 121 |  |
| Hyperlipidemia | Yes | 456(28.7%) | 736(35.3%) | 0.130 | 150 | 152 | 0.996 |
|  | No | 1,135(71.3%) | 1,351(64.7%) |  | 318 | 322 |  |
| Diabetes mellitus | Yes | 585(36.8%) | 890(42.6%) | 0.209 | 191 | 203 | 0.513 |
|  | No | 1,006(63.2%) | 1,197(57.4%) |  | 278 | 271 |  |
| Atrial fibrillation | Yes | 140(8.8%) | 176(8.4%) | 0.933 | 45 | 42 | 0.697 |
|  | No | 1,451(91.2%) | 1,911(91.6%) |  | 423 | 431 |  |
| Previous stroke | Yes | 379(23.8%) | 302(14.5%) | 0.009 | 89 | 88 | 0.859 |
|  | No | 1,212(76.2%) | 1,785(85.5%) |  | 379 | 386 |  |
| MMSE score | 2 | 84(5.3%) | 77(3.7%) | 0.591 | 21 | 21 | 0.989 |
|  | 3 | 254(16.0%) | 302(14.5%) |  | 67 | 66 |  |
|  | 4 | 1,253(78.7%) | 1,708(81.8%) |  | 381 | 386 |  |

Continuous variables are presented as mean ± standard deviation, and categorized variables are presented as n (%).
PAC, post-acute care; MMSE, Mini-Mental State Examination.

### Structural Equation Modeling of Overall Depression or Anxiety

The participation of PAC positively predicted cognitive function at week 12 (b=1.42, P<0.001) (Figure 1). This subsequently influenced patients’ depression scores at week 12 and at year 1. Particularly, pre-rehabilitation (baseline) cognitive function positively predicted depression scores at week 12 (b=0.05, P<0.001). However, whereas the path of PAC on cognitive function at year 1 was not significant but had a direct effect on depression at year 1 (b=0.88, P<0.001). Cognitive function at week 12 fully mediated between PAC participation and depression at week 12 (b=-0.07, P<0.001). At year 1, the participation of PAC had a direct effect on depression. Furthermore, at week 12, cognitive function fully mediated between PAC participation and anxiety at week 12 (b= -0.07, P<0.001) (Figure 2). However, whereas at year 1, PAC participation had a direct impact on anxiety (b=1.01, P<0.001). Overall, besides the mediating effect of the depression-anxiety model, there were positive prediction effects among cognitive function, depression, and anxiety at different assessment time points, with all paths being significant (P<0.05). In Figure 1, the pre-rehabilitation cognitive function affected cognitive function at week 12 (b=0.76, P<0.001), and the cognitive function at week 12 affected the cognitive function at year 1 (b=0.85, P<0.001). Regarding depression, pre-rehabilitation depression affected depression at week 12 (b=0.18, P<0.001), and depression at week 12 also affected depression at year 1 (b=0.19, P<0.001). In Figure 2, regarding anxiety, pre-rehabilitation anxiety affected anxiety at week 12 (b=0.15, P<0.001). Anxiety at week 12 also affected anxiety at year 1 (b= 0.20, P<0.001). These findings suggest that the patient’s cognitive function, depression, and anxiety were affected over time.

**FIGURE 1.**
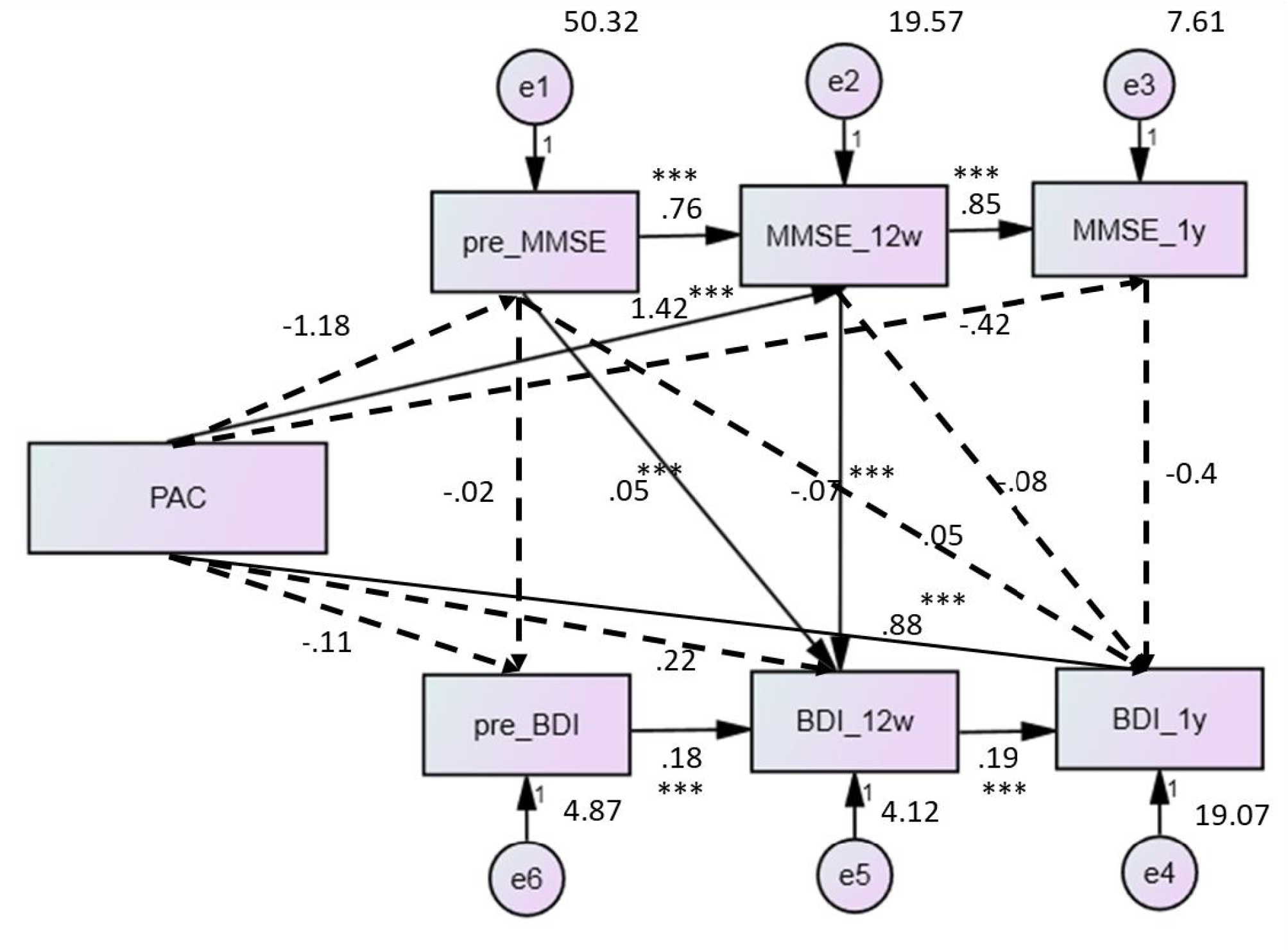
The whole model using structural equation model. The solid line represents a significant relationship between the two variables. Numbers are path coefficients. PAC, post-acute care; MMSE, mini-mental state examination; BDI, Beck Depression Inventory; BAI, Beck Anxiety Inventory. *P<0.05, **P<0.01, ***P<0.001. To investigate whether cognitive function mediates the effect of enrollment in PAC programs on depression in stroke patients and whether the indicators are moderated in the pathway;

**FIGURE 2.**
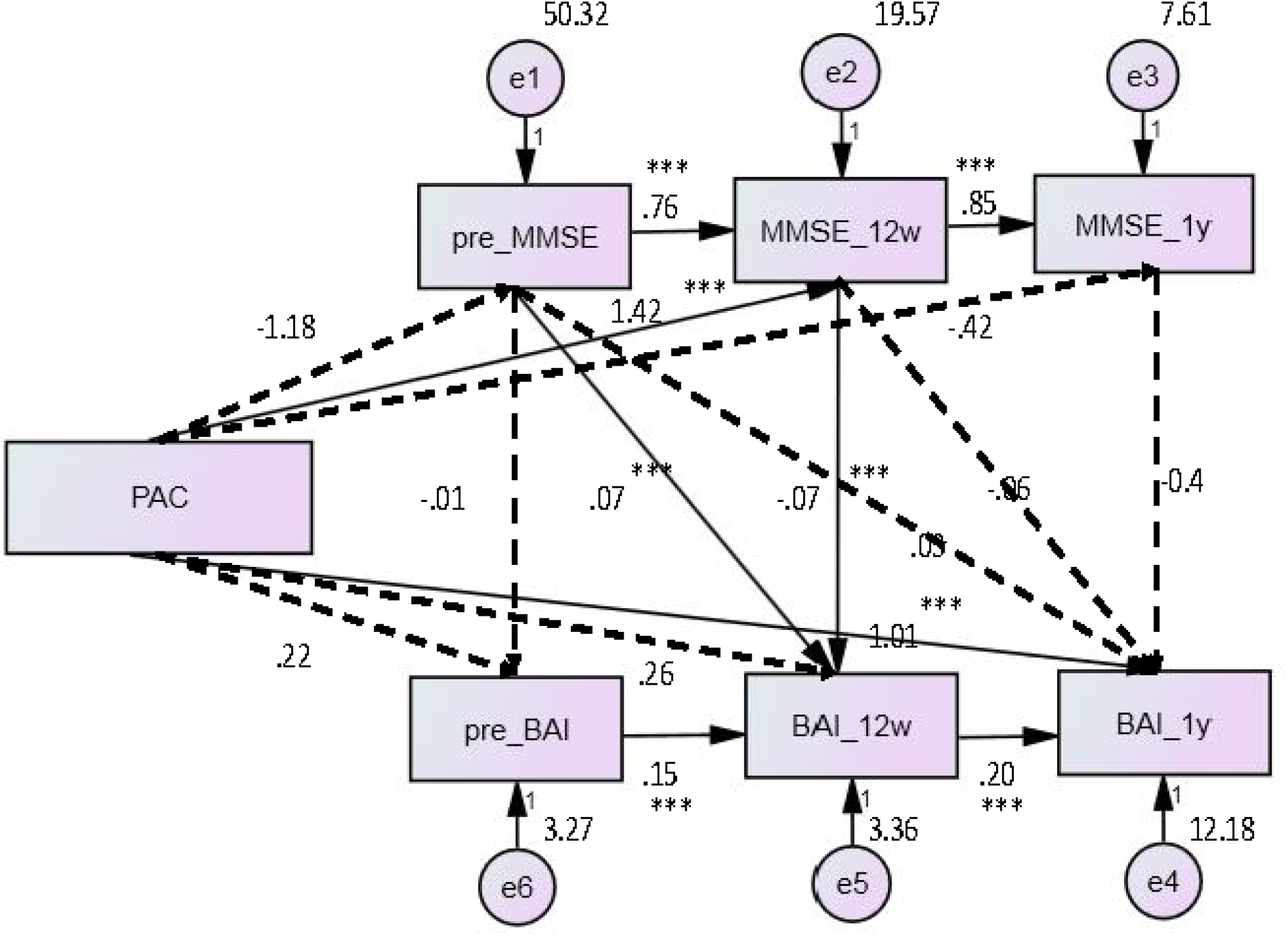
The whole model using structural equation model. The solid line represents a significant relationship between the two variables. Numbers are path coefficients. PAC, post-acute care; MMSE, mini-mental state examination; BDI, Beck Depression Inventory; BAI, Beck Anxiety Inventory. *P<0.05, **P<0.01, ***P<0.001. To investigate whether cognitive function mediates the effect of enrollment in PAC programs on anxiety in stroke patients and whether the indicators are moderated in the pathway.

### Moderating Effect

First, in the path from the independent variables to the mediating variables of the 12^th^ week depression model (education (b=0.582) and acute lengths of stay (b=0.203)) and anxiety model (education (b=0.517) and acute lengths of stay (b=0.131)), had a moderating effect (P<0.05) (Table 2). Furthermore, in the path from the mediating variables to the dependent variables of the 12^th^ week depression model (age (b=-0.002), alcohol drinking (b=-0.145), acute lengths of stay (b=-0.003), foley catheter (b=-0.112), nasogastric tube (b=-0.629), and diabetes mellitus (b=-0.647)) and anxiety model (body mass index (b=0.006), acute lengths of stay (b=-0.004), and previous stroke (b=-0.092)) had a significant moderating effect (P<0.05) (Table 3). Additionally, the results in Tables 2 and 3 indicate that acute lengths of stay as a moderating variable was significant in both paths of the depression and anxiety models. Therefore, acute lengths of stay was added as a moderating variable in the mediating and moderating effects analysis.

**Table 2.** The Moderation Effect in the Path from the Independent Variables to the Mediating Variables of the 12^th^ Week Depression or Anxiety Model After Post-Acute Care (PAC) Program.

| Moderator (W) | Depression Model |  | Anxiety Model |  |
| --- | --- | --- | --- | --- |
|  | b | P value | b | P value |
| Age, years | -0.092 | 0.051 | -0.081 | 0.059 |
| Gender, female vs. male | 0.752 | 0.579 | 0.827 | 0.599 |
| Body mass index, kg/m<sup>2</sup> | 0.123 | 0.488 | 0.130 | 0.540 |
| Education, years | 0.582 | 0.024 | 0.517 | 0.020 |
| Smoking, yes vs. no | 0.628 | 0.686 | 0.670 | 0.699 |
| Drinking wine, yes vs. no | -0.394 | 0.841 | -0.434 | 0.887 |
| Acute lengths of stay, days | 0.203 | 0.002 | 0.131 | 0.001 |
| Readmission in 30 days, yes vs. no | 0.904 | 0.714 | 0.947 | 0.841 |
| Stroke recurrence, yes vs. no | -2.269 | 0.456 | -2.137 | 0.407 |
| Foley catheter, yes vs. no | 1.818 | 0.380 | 1.751 | 0.307 |
| Nasogastric tube, yes vs. no | -0.489 | 0.740 | -0.402 | 0.674 |
| Hypertension, yes vs. no | -0.414 | 0.775 | -0.497 | 0.798 |
| Diabetes mellitus, yes vs. no | 1.206 | 0.366 | 1.362 | 0.398 |
| Hyperlipidemia, yes vs. no | 1.442 | 0.305 | 1.167 | 0.275 |
| Atrial fibrillation, yes vs. no | 1.501 | 0.517 | 1.576 | 0.501 |
| Previous stroke, yes vs. no | 1.419 | 0.394 | 1.294 | 0.300 |
| MMSE scores | 0.400 | 0.739 | 0.412 | 0.727 |

MMSE, Mini-Mental State Examination.

**Table 3.** The Moderation Effect in the Path from the Mediating Variables to the Dependent Variables of the 12^th^ Week Depression or Anxiety Model After Post-Acute Care (PAC) Program.

| Moderator (W) | Depression Model |  | Anxiety Model |  |
| --- | --- | --- | --- | --- |
|  | b | P value | b | P value |
| Age, years | -0.002 | 0.028 | -0.001 | 0.366 |
| Gender, female vs. male | 0.036 | 0.224 | -0.037 | 0.162 |
| Body mass index, kg/m<sup>2</sup> | -0.001 | 0.693 | 0.006 | 0.050 |
| Education, years | -0.001 | 0.953 | -0.006 | 0.069 |
| Smoking, yes vs. no | -0.016 | 0.626 | 0.018 | 0.553 |
| Drinking wine, yes vs. no | -0.145 | <0.001 | -0.042 | 0.270 |
| Acute lengths of stay, days | -0.003 | 0.021 | -0.004 | 0.002 |
| Readmission in 30 days, yes vs. no | 0.054 | 0.296 | 0.042 | 0.375 |
| Stroke recurrence, yes vs. no | 0.046 | 0.354 | -0.006 | 0.897 |
| Foley catheter, yes vs. no | -0.112 | 0.009 | -0.009 | 0.812 |
| Nasogastric tube, yes vs. no | -0.629 | 0.039 | -0.018 | 0.511 |
| Hypertension, yes vs. no | -0.430 | 0.219 | 0.054 | 0.087 |
| Diabetes mellitus, yes vs. no | -0.647 | 0.019 | -0.009 | 0.714 |
| Hyperlipidemia, yes vs. no | -0.001 | 0.981 | 0.005 | 0.850 |
| Atrial fibrillation, yes vs. no | 0.049 | 0.381 | -0.065 | 0.198 |
| Previous stroke, yes vs. no | -0.011 | 0.758 | -0.092 | 0.005 |
| MMSE scores | 0.005 | 0.900 | -0.012 | 0.733 |

MMSE, Mini-Mental State Examination.

### Analysis of Mediating and Moderating Effects

Regarding the depression and anxiety mediating and moderating models, PAC significantly predicted cognitive function at week 12 (a1=-2.617, P=0.043) (Table 4). Acute lengths of stay significantly predicted cognitive function at week 12 (a2=-0.214, P<0.001). The interaction between PAC and acute lengths of stay significantly predicted cognitive function at week 12 (a3=0.204, P=0.002). In the mediating and moderating models, cognitive function at week 12 did not significantly predict depression and anxiety at week 12. In contrast, acute lengths of stay significantly predicted depression and anxiety at week 12 (b2=0.074, P=0.015 and b2=0.094, P=0.001, respectively). The interaction between cognitive function at week 12 and acute lengths of stay also significantly predicted depression and anxiety at week 12 (b3=-0.003, P=0.021 and b3=-0.004, P=0.002, respectively). Additionally, PAC positively influenced the direct effect of depression and anxiety at week 12 but not at a significant level. This indicates that although acute lengths of stay had a significant moderating effect, the original mediating effect of the depression and anxiety models was lost when the moderating effect was added.

**Table 4.**
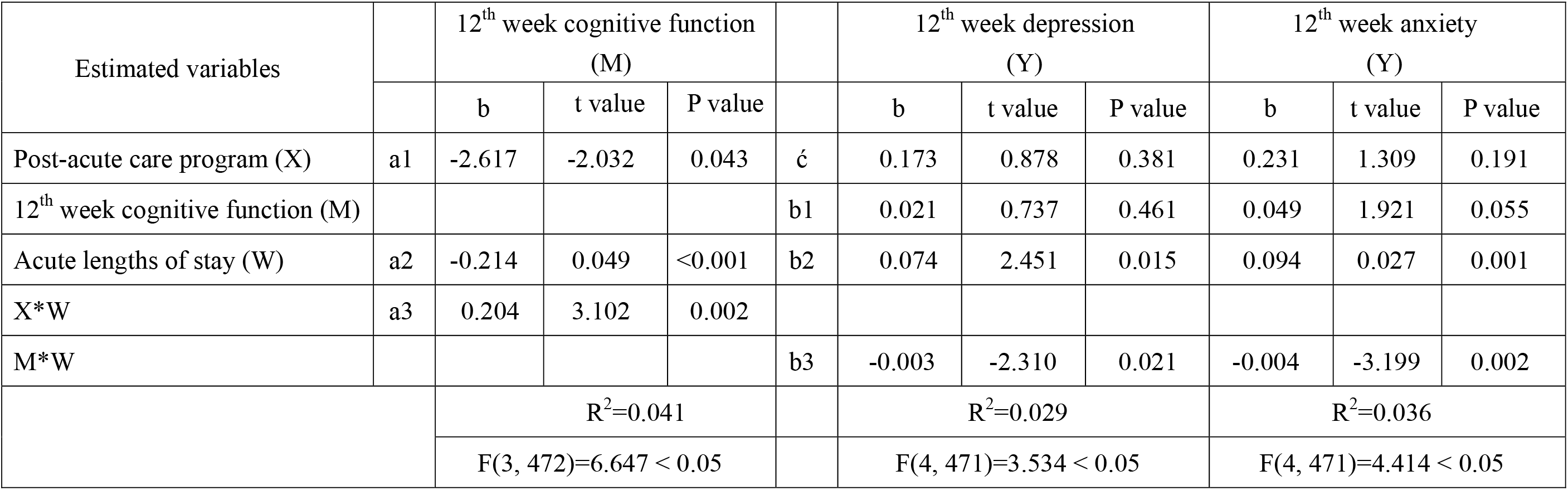
The Mediation Effect of Depression and Anxiety and the Moderation Effect on All Paths.

## DISCUSSION

Although PAC has been promoted for the past decades, evidence of its mediating effect of cognitive function has been limited.^15, 16^ To address this issue, we used IPTW matching in this study. It has been demonstrated that there was a significant improvement in cognitive function at week 12 in the PAC group compared to the non-PAC group (P<0.05), but there was no significant difference at year 1.^17^ It also has noted significant improvements in cognitive function scores after a mean of 43.57 days of PAC participation,^18^ which is consistent with the path significance in the current study. There was no correlation between PAC participation and cognitive function in the long term, but there was a more significant correlation in the short term. One study was designed to examine how long that cognitive improvement lasts and to compare depressed patients’ cognitive status with that of non-depressed patients with comparable lesions.^19^ They concluded that stroke patients who receive treatment consistently in the early stages of depression can sustain cognitive improvement for more than two years. Therefore, providing intensive rehabilitation therapy early in the progression course can help improve patients’ depression symptoms.

The present study is consistent with previous literatures in that anxiety symptoms after stroke were associated with cognitive function, physical function, and quality of life.^20–22^ Moreover, anxiety symptoms after stroke were significantly associated with lower cognitive function before rehabilitation. However, no significant correlation was observed between the vertical association of anxiety symptoms after stroke with changes in cognitive function over one year.^23, 24^

Education had a moderating effect on the PAC’s prediction of cognitive function at week 12. Age and alcohol drinking had a significant moderating effect on cognitive function at week 12 for predicting depression at week 12. However, it had no moderating effect on cognitive function at week 12 for predicting anxiety at week 12. It has suggested that education is associated with cognitive function and that those with higher education had significantly better cognitive performance than those with lower education.^25, 26^ In the present study, at three months after stroke, 37% of the patients were at risk for depressive symptoms. Further, depression scores at three months after stroke were higher in patients aged 25-54 years than in those aged 75 years and older, as well as in those aged 55-64.^27^ Past studies indicated that alcohol drinking was significantly associated with the severity of depressive symptoms and that depressive symptoms were more severe in women than men.^28, 29^ PAC was not a significant predictor of cognitive function at week 12. In contrast, a nasogastric tube, a foley catheter, and diabetes mellitus were significant moderators in the path of cognitive function at week 12, predicting depression at week 12. Previous stroke was a significant moderator in the path of cognitive function at week 12, predicting anxiety at week 12. Therefore, depressive symptoms will increase the risk of developing diabetes mellitus by 1.62 to 2.52 times, suggesting that the product (interaction) of diabetes mellitus and cognitive function also predicts depression.^30, 31^ Another study found a significant association between severe anxiety symptoms and an increased risk of stroke.^32^ However, age, education, alcohol drinking, a nasogastric tube, a foley catheter, diabetes mellitus, and previous stroke had only moderating effects in a single path, not in both paths. Thus, demographic attributes and clinical attributes were not included in the final mediating and moderating effects analysis of the present study.

Acute lengths of stay was significant in each model path for depression and anxiety symptoms but was not significant for stroke recurrence and readmission in 30 days. First, the average lengths of stay for the patients with stroke in Taiwan was longer than it in foreign countries.^3, 33^ It also indicated that acute lengths of stay was negatively associated with depression and anxiety symptoms after one month’s hospital treatment.^34^ In contrast, readmission in 30 days was not associated with depression and anxiety symptoms. The mediating model lost its mediating effect when acute lengths of stay was added as a moderating variable; the week 12 cognitive function predicting week 12 depression and the path of depression was not significant. This could be explained as the mediating effect of acute lengths of stay in the PAC prediction of the week 12 cognitive function path. Acute lengths of stay was significantly correlated to post-rehabilitation cognitive function and was an independent predictor, and could have a more significant effect as a mediating variable in the model than as a moderating variable.

There are three limitations to the current study. First, we only included stroke patients from six hospitals in Taiwan, which is not representative of patients with stroke across Taiwan or other regions and is regionally limited. Furthermore, the follow-up period was one year, which lacks long-term follow-up of patients and cannot investigate the long-term effects of PAC or changes in cognitive function and psychological status. Finally, the patients dictated the questionnaire, and interviews were conducted in multiple stages, which may be influenced by patients’ subjective judgment, recall bias, or social expectations.

PAC improved the anxiety and depression symptoms of patients with stroke after 12 weeks via the effects of cognitive function and directly affected anxiety and depression symptoms after one year of rehabilitation. These findings suggest that participation in PAC program can improve anxiety and depression symptoms in stroke patients. The cognitive function, anxiety, and depression symptoms of patients with stroke who participated in PAC program changed over time after rehabilitation, i.e., the earlier the patients received rehabilitation, the more their anxiety and depression symptoms improved. Thus, cognitive function mediated the relationship between PAC and depression and anxiety symptoms. Other moderating variables influenced it, while the mediating role may be lost with the addition of moderation. Therefore, we recommend that healthcare providers should give multiple cognitive rehabilitation or psychotherapy to the long-term inpatients participating in PAC.

## Data Availability

The datasets used/or analyzed during the current study are available from the corresponding author on reasonable request.

